# Burden of predominant psychological reactions among the healthcare workers and general during COVID-19 pandemic phase: a systematic review and meta-analysis

**DOI:** 10.1101/2021.01.02.21249126

**Authors:** Bhaskar Thakur, Mona Pathak

## Abstract

**Aim:** Present systematic review and meta-analysis examined the burden of psychological reactions predominantly anxiety, depression, stress and insomnia during novel COVID-19 pandemic phase among the frontline healthcare, non-frontline healthcare and general.

**Methodology:** PubMed, EMBASE and SCOPUS were searched for studies between Jan 1, 2020 to May 25, 2020. Brief protocol of the systematic review was registered with the PROSPERO database, (CRD42020186229).Any study that reported the burden of at least one of psychological reactions including anxiety or depression or stress or insomnia was eligible. Heterogeneity was assessed using I^2^ statistic and results were synthesized using random effect meta-analysis.

**Results:** Out of 52eligible studies, 43 reported anxiety, 43 reported depression, 20 reported stress and 11 reported insomnia. Overall prevalence for anxiety, depression, stress and insomnia were 26.6%, 26.2%,26.2% and 34.4% respectively. Anxiety and depression were found highest among the COVID-19 patients (43.3% and 51.75 respectively). Apart from COVID-19 patients, prevalence of anxiety, depression, stress and insomnia were found highest among the frontline healthcare (27.2%, 32.1%,55.6% and 34.4% respectively) as compared to general healthcare workers (26.9%, 15.7%, 7.0% and 34.0% respectively) and general population (25.9%, 25.9%,25.4% and 29.4% respectively).

**Conclusion:** Anxiety and depression were found highest among the COVID-19 patients. Apart from COVID-19 patients, the anxiety, depression, stress and insomnia were more prevalent among frontline healthcare workers compared to general. Such increased prevalence is prompting towards the global mental health emergency. Therefore a call of urgent attention and pan-region effective mental-health intervention are required to mitigate these psychological reactions.

## 1. Introduction

The novel pneumonia caused by the coronavirus disease (COVID-19) has emerged in the Chinese city of Wuhan (Hubei province) in late December 2019 and spread rapidly nationwide and all over the world(1,2). The World Health Organization (WHO) called an emergency meeting on COVID-19 and declared an international public health emergency on January 30, 2020. The virus spread in nearly 213 countries and territories with 66.2 million confirm cases, 1.5 million confirmed deaths and 45.8 million recovered according to the global data reported by the Worldometer on Dec 4, 2020(3).

During this global pandemic fear of rapid infection spread, falling sick and dying, social isolation and extended quarantine are expected to influence the mental health. Fear related to unavailability of treatment and vaccine, shortages of critical care support and other medical equipment’s as well as sudden shortfall in mask and hand sanitizers in the market may result into psychological distress and other mental health illness. Additionally, fear of financial crisis, joblessness, and frozen economy, during lockdown may play the lead role to increase the burden of mental health illness. Such psychological burden had been reported among the COVID-19 patients, healthcare personnel, medical students, and older as well as general population. (2,4–6)

Mental-illness was globally estimated around 32.4% of total year lived with disability and 13.0% of disability adjusted life years.(7) Depression was the first leading cause of disability and a prime contributor to the disease burden globally (8) whereas anxiety disorders reported as sixth leading causes of global disability. (9) Sudden declaration of public health emergency may further intensify the existing burdens of mental health outcomes. History has been witnessing to the mental health challenges during infectious outbreaks around the globe. (10) During Ebola outbreak in West Africa, greater number of healthy people was mentally traumatized compare to the number of infected people, and remained longer (11). Such historical devastation prompts towards another global mental health challenges during COVID-19 pandemic. Therefore, it is important to understand the increased burdens of mental health outcomes as a consequence of COVID-19 pandemic.

Several studies suggested that the new psychological reactions were uncovered during initial phase of COVID-19 pandemic. However, estimates of these burdens vary across the studies. Such variations might occur because these studies carried on different population, with varying sample sizes and dealt with different scale of mental-illness assessment.

### 1.1 Aim of the study

In this study, a systematic review and meta-analysis was conducted on the burden of mental health outcomes predominantly on the prevalence of anxiety, depression, stress and insomnia during COVID-19 global emergency. We focused to assess the burden among three group of population, i.e., frontline healthcare workers (FHW), non-frontline healthcare workers (NFHW) and general who are not healthcare workers.

## 2. Material and Methods

Prior to conducting this systematic review and meta-analysis, the study aims and methods (brief protocol) were registered with the PROSPERO database, University of York (Registration Number: CRD42020186229)(12). The present systematic review manuscript is designed as per the guidelines of Preferred Reporting Items for Systematic Reviews and Meta-Analysis (PRISMA) (13).

### 2.1 Data Source and Search strategy

A systematic literature search in electronic databases including Embase, PubMed and Scopus between January 1, 2020 and May 25, 2020 was used in order to find the eligible studies. The used search term was”(COVID-19 OR SARS-CoV-2 OR 2019-nCoV OR corona virus) AND (Depression OR Anxiety OR stress OR insomnia OR “psychological distress” OR “Psychiatric illness” OR “Mental Health”)”. Additionally, a supplementary search was conducted using Google Scholar.

### 2.2 Eligibility criteria

All the published or unpublished studies were considered eligible if they met the following eligible criteria: (1) described the assessment of at least one of mental-illness including psychological anxiety or depression or stress or insomnia as an impact of COVID-19; (2) used the scientific rating scale to assess the mental-illness; and (3) the scale based findings were reported in terms of overall prevalence and graded prevalence (mild, moderate and severe). All those studies published as letter to editor, editorial or commentaries and reported the required outcome were also included.

### 2.3 Study Selection& Data Extraction

All the retrieved articles first were screened on the basis of title and abstract and then reviewed for full text of potentially eligible articles independently and in duplicate by both authors (B.T. and M.P.). Data regarding study identification, population, sample size, prevalence of anxiety, Depression, Stress and Insomnia (or categorized on graded scale like normal, mild, moderate, and severe), scale for outcome measurement and quality related variables were extracted by both authors independently on pre-prepared form. All the discrepancies were resolved by discussion.

### 2.4 Methodological quality assessment

The modified Newcastle-Ottawa Quality Assessment Scale (NOS) was used to assess the methodological quality for cross-sectional studies (14) (Supplementary Figure 1). Since present meta-analysis focused only prevalence of mental health outcomes, the quality of the studies was judged on the basis of four criteria only including representative sample, adequate sample size, low non-response rate and objective outcome measurement. So the range of quality score was 0 to 5. Quality scores were categorized as 4/5= Good, 3= Average, 2/0= Poor. Both the reviewer (BT and MP) independently assessed the quality of eligible studies.

### 2.5 Summary Measures& Synthesis

Proportion of psychological stress, anxiety, depression and insomnia as overall, as well as on the graded scale of mild, moderate and severe were pooled using fixed effect inverse variance method or DerSimonian& Liard random effect method(15,16) depending on heterogeneity measured using *I*^*2*^ statistic (17). Subgroup analysis was performed on the basis of type of population, i.e., FHW, NFHW and other general.

Publication bias assessment method lays on the fact that likelihood of publication depends on the sample size and statistical significance (18), but this was not the case with the current research conducted during Covid-19. Since conventional funnel plots are inaccurate to assess the publication bias in meta-analysis of proportion, especially in case of proportion (19), we used egger’s test (20) to assess publication bias. The quality of our evidence was graded using GRADE (Grading of Recommendations, Assessment, Development and Evaluations) approach(21).

## 3. Results

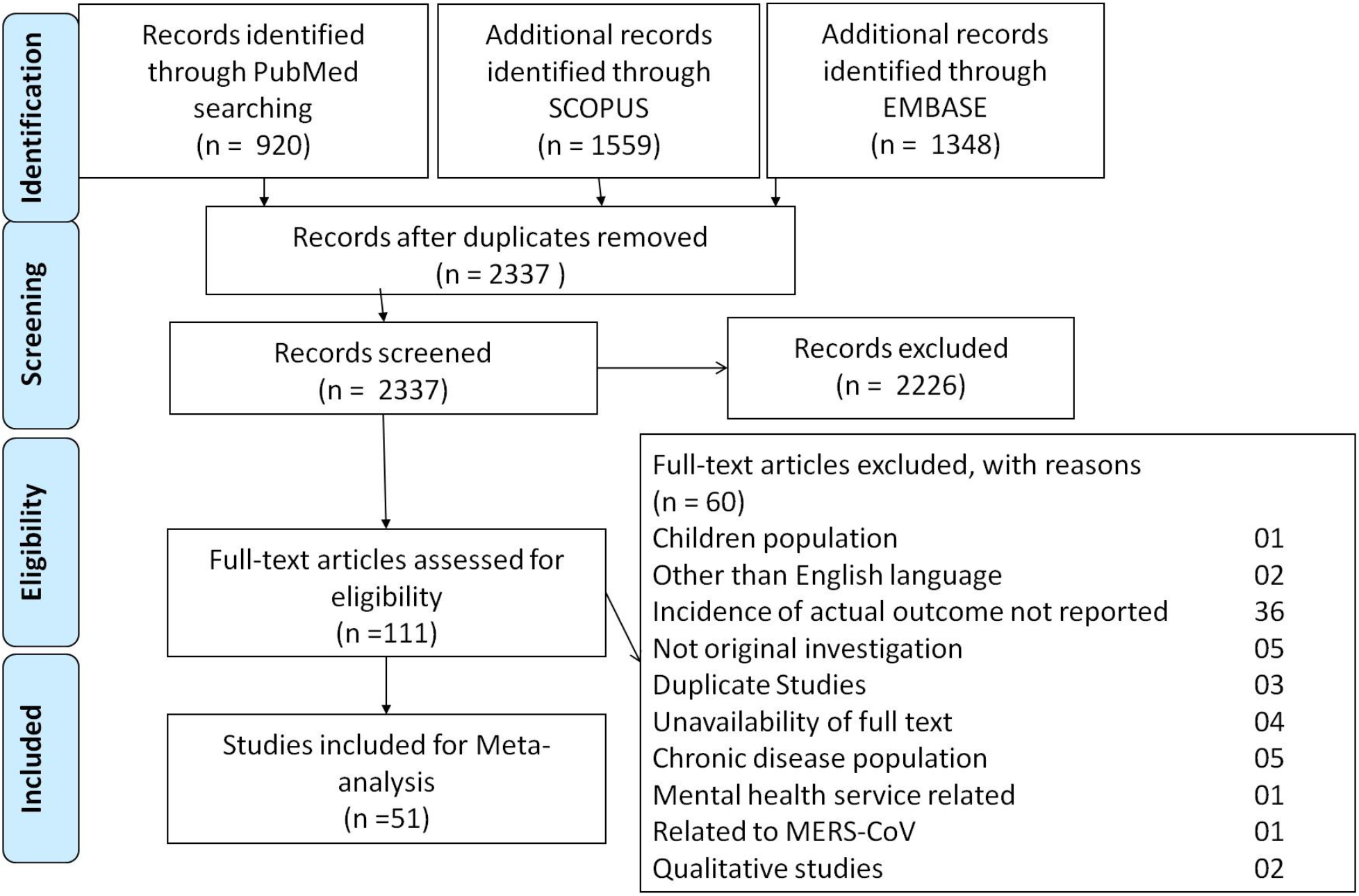

### 3.1 Study Selection

A total of 2337 unique studies were identified by searching the databases. Out of these, 111 studies qualified for full text review [Figure 1]. Out of these 111 studies, 60 were excluded because of various reasons such as one study with children population, two studies found published in other than English language, incidence of actual outcome not reported in 36 studies, original investigation were not done in five studies, three studies were found duplicate, full text was not available for four studies, five studies found in chronic disease population, one study was related to mental health service, one study found related to MERS-CoV and two were qualitative studies.

### 3.2 Study Characteristics

Our search yielded a total of 51 studies for inclusion which comprised 59 datasets reported various types of mental health among diverse population for 147142 individuals. Out of these 59 datasets, 32 datasets involving 123650 individuals reported mental health burden for general population(5,22–52); 15 datasets from 14 studies reported mental health of 8335 frontline healthcare workers(35,52–65); eight datasets from seven studies reported for 12462 non-frontline health care workers(4,28,39,66–69), two datasets represented 160 COVID-19 Patients(49,70) and two datasets reported mental health of 2535 quarantined populations(49,71).

Two studies(35,52) reported mental health status of FHWs as well as general population while another two studies(28,39) reported among the healthcare workers other than frontline as well as general population. One study(49) reported mental health incidences independently among the COVID-19 patients, quarantined group and general population. One study (56) reported anxiety and depression among the FHWs with different sample sizes. Group-wise data in these studies were extracted to facilitate subgroup comparison.

Majority of the studies (35 out of 52) were conducted in China. Among rest of the 17 studies, two (29,37) were conducted in Iran, one was in Israel (68), one study was conducted in Singapore (66), three were in Spain(26,40,41), one study in Greece (46), one study from France(53), one study from USA(30), two study from Turkey (24,42), one from England (56) and four studies were conducted in Italy (23,36,54,67). Further, the study conducted in Singapore (66) included data for India as well as Singapore. So country level data was extracted for this study. Most of the studies used online surveys to collect data.

The severity of psychological health parameters were reported by 19 studies (4,5,22,27,31,37,41,45,47,51,55,56,58,59,61,62,64,67,70); 18 for anxiety (4,5,22,27,31,37,41,45,47,51,55,56,58,59,61,64,67,70); 13 for depressio (5,22,27,31,41,45,47,51,56,59,64,70,71); five for stress(5,27,41,45,59) and four for insomnia (27,45,59,62). Details of study level characteristics are reported in supplementary Table 1.

### 3.3 Quality assessment

Out of the total 51 included studies, 32 studies had ‘Good’ quality (12 studies had score 5 and 20 studies had score 4), 18studies were grades as ‘average’ quality with score 3 and one study had poor quality with score 2.Details of quality assessment for these included studies are made available in supplementary Table 2. Majority of the studies collected data from online survey on social media platform. These studies could include data for social media users not from any well-defined population and hence these studies did not have representative sample. Formal sample size calculation or power assessment was done only for two studies. Further we found a wide heterogeneity in the reported results. These facts lower the confidence in our graded evidence. The moderate grade for evidence of anxiety and depression suggests that further studies may less likely to change the current evidence. Assessment of Confidence in our reported finding by GRADE approach are made available in supplementary Table 3.We could not find significant publication bias for any of the outcome.

### 3.4 Anxiety outcome

Overall pooled prevalence of anxiety was 26.6% (95% CI: 22.8% – 30.4%) which was observed relatively higher among the COVID-19 patients (43.3%; 95% CI: 36.0% - 50.7%)followed by FHWs (27.2%; 95% CI: 18.1% – 36.3%), NFHW(26.9%; 95% CI: 20.3% - 33.4%), general population (25.9%; 95% CI: 20.5% – 31.2%), and quarantined group (10%; 95% CI: 4.4% - 21.4%)[Figure 2 (A)].

Meta-analysis of prevalence on severity scale based on 18available studies resulted that most of the individuals had mild anxiety (16.7%; 95% CI: 12.3% - 21.1%). Moderate anxiety (7.3%; 95% CI: 4.4%- 10.3%) and severe anxiety (5.4%; 95% CI: 3.2%-7.6%) were observed among very few individuals [Figure 3 (A)]. However, moderate and severe anxiety was observed highest among the general population. Figure 3(A) clearly demonstrated the anxiety burden on severity scale in different general.

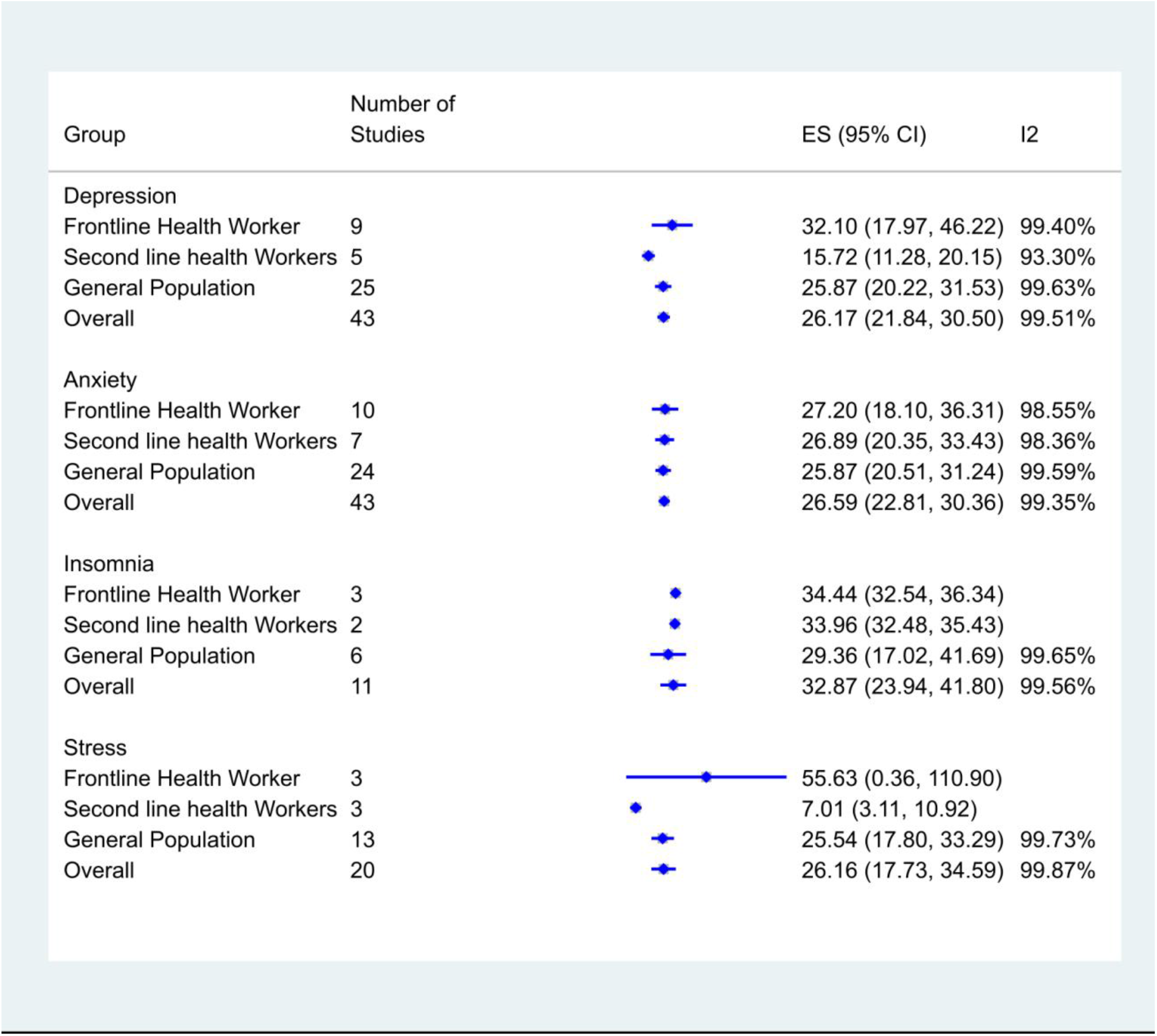

### 3.5 Depression outcome

A total of 26.2% (95% CI: 21.8% - 30.5%) were identified to have depression (Figure 2(B)). This prevalence was found highest among the COVID-19 patients (51.7%; 95% CI: 44.2% - 59.3%) followed by the FHWs (32.1%; 95% CI: 18.0% - 46.2%), general population (25.9%; 95% CI: 20.2% - 31.5%), NFHW(15.7%; 95% CI: 11.3% - 20.1%) and quarantine group (9.1%; 95% CI: 7.9% - 10.2%)[Figure 2(B)].On severity scale of depression, 21% of the individuals had mild depression, 7.4% had moderate and very few (4.6%) were severe cases [Figure 3(B)]. Further, depression burden was observed highest among the frontline health workers in all the three group of mild, moderate and severe [Figure 3(B)].

### 3.6 Stress assessment

Figure 2(C) showed the overall stress prevalence as 25.1% (95% CI: 16.9% – 33.3%). Our pooled effect of stress prevalence was found highest among the FHWs (55.6%; 95% CI: 0.36% – 110.9%) followed by general population (23.9%; 95% CI: 15.9% – 32.0%), NFHW (7.0 %; 95% CI: 3.1% - 10.9%) and quarantined group with single study (2.7%; 95% CI: 2.1% - 3.4%). Based on five reported studies, pooled stress prevalence resulted into 18.3% mild, 9.8% moderate and only 4.4% severe stress levels [Figure 3(C)].

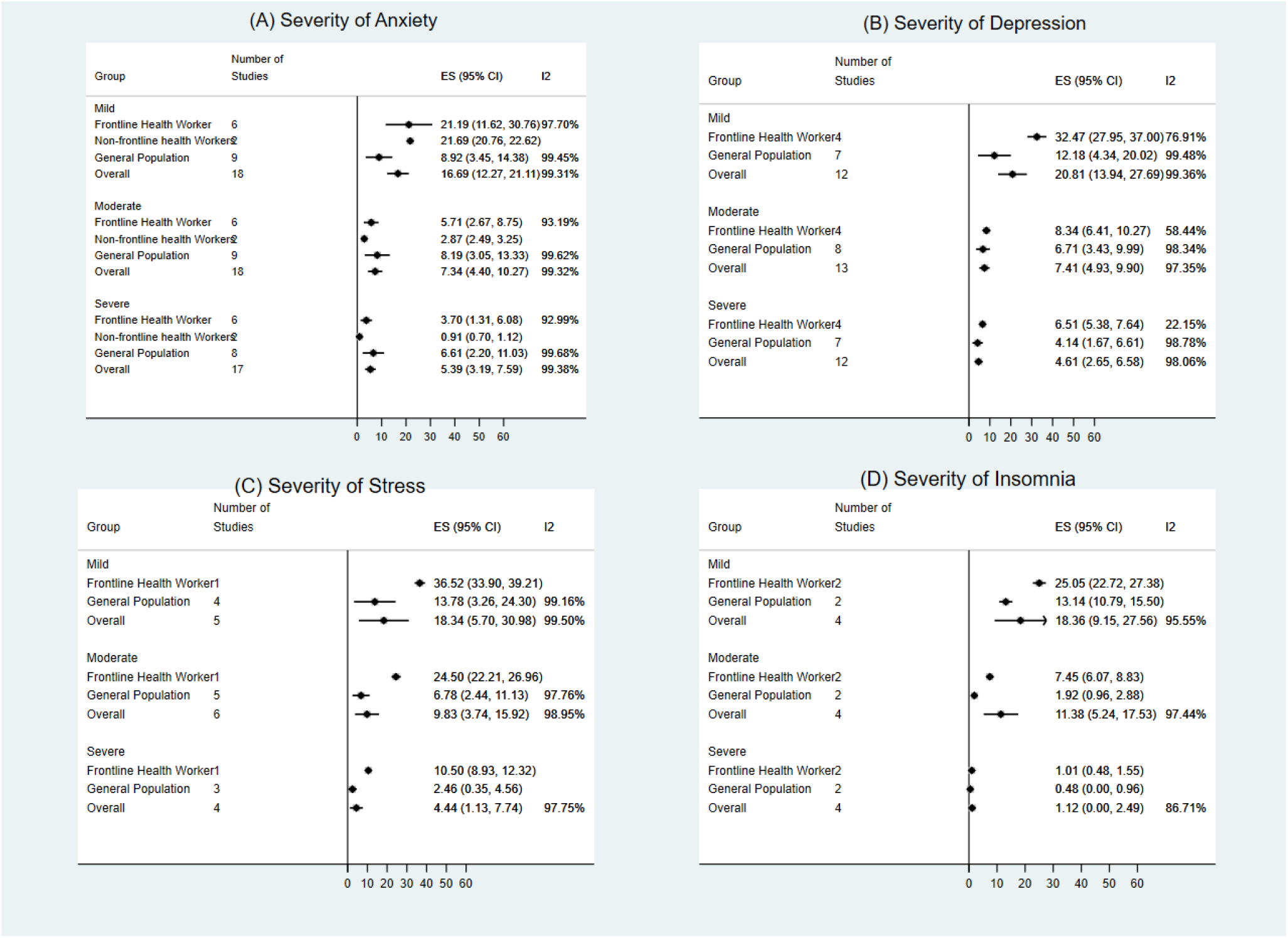

### 3.7 Insomnia assessment

Overall insomnia burden was found as 31.3% (95% CI: 22.9% - 39.7%) [Figure 2(D)]. Pooled effect of insomnia prevalence among the general population was observed as 27.2% (95% CI: 16.3% - 38.2%), a lower than the prevalence among FHWs (34.4%; 95% CI: 32.5% - 36.3%) and NFHWs (34.0%; 95% CI: 32.5% - 35.4%) [Figure 2(D)]. Further on the severity graded scale, mild, moderate and severe level of insomnia were observed as 18.4%, 11.4% and 1.1% respectively [Figure 3(D)].

## 4. Discussion

### 4.1 Summary and evidence

This systematic review and meta-analysis was based on the data extracted from 51 different studies. Most of these studies were conducted in China, the country where COVID-19 emerged, although studies conducted in Taiwan, Vietnam, Singapore, Italy, Israel, Iran, Greece, Spain, USA, Turkey and England were also included. Most of the studies were conducted using online platform of social media. Studies varied in sample size from 57 to 52730. One largest study (43) incorporated information for multiple countries like China, Macau, Hong Kong and Taiwan. A total of 43 studies reported prevalence of anxiety and depression. Stress and insomnia were reported by few studies, i.e., 21 and 12respectively. Various scales were used to measure these outcomes but most of the variability found for stress outcome. All the included studies were performed with cross-sectional design and majority of these studies were based on web-based survey and therefore lacking random sampling method of data collection. Grade approach suggested there is moderate level of confidence in our finding for anxiety, depression and insomnia but low level of confidence for stress.

Burden of anxiety and depression in COVID-19 patients was found the highest compared to general. However these results were based on only two available studies. Apart from COVID-19 patients, burden of anxiety, depression, stress and insomnia were found highest among the FHWs. We also found that after FHWs, burden of depression and stress was highest among the general population and burden of anxiety and insomnia was highest among the NFHWs.

During this pandemic, a handful of reviews and meta-analysis on the prevalence of mental health outcomes were reported among the healthcare workers and general population. (44,72,73) However, all these reviews are based on small number of studies, majorly based on Chinese studies and focusing a particular population. In addition to the Chinese studies, our review attempted to include the most updated global studies targeting wide range of population. We also attempted the overall prevalence based on the graded scale of severity for all the outcomes. Burden of mild stage were observed as highest followed by moderate and severe stages for all the four outcomes. Similar exploration on graded scale was also attempted among the various subpopulations, i.e., FHWs, NFHWs, general population, COVID-19 infected patients and quarantine people.

A meta-analysis reported that the global prevalence of anxiety disorder was 7.3% in general population after adjusting the methodological differences.(74) According to the recent report of *Our World in Data*, prevalence of anxiety disorder and depression was observed as 3.8% and 3.4% respectively. (75) Our finding suggests how the psychological pressure during the pandemic public health crisis increased the mental health burden. According to a recent meta-analysis report in 2017(76), insomnia prevalence in the general population of China observed as 15% which was far lower than the insomnia prevalence among the general population observed in our study during COVID-19 pandemic. Similarly the global prevalence of post-traumatic stress disorder was reported as 15.3%.(77) These reports and our finding suggest that the burden of insomnia and stress in general population, increased almost to double, was certainly a consequences of COVID-19 fear.

During the crucial public health emergency of COVID-19, the frontlines healthcare professionals feel fear of getting sick and spreading infection to their families, other patients and coworkers. They face potential mental health crisis with either sudden shortages of personal protective equipment (PPE) at their workplace or when they get positive with COVID-19 test. Apart from the COVID-19 infected patients, our comparative analysis results shows that the prevalence of all four mental health outcomes was significantly high among the frontline healthcare professionals as compared to general. These findings suggest the positive correlation between management of COVID-19 patients by healthcare professionals and increased psychological responses among them.

Although surveys and studies in current COVID-19 emergency confirmed new psychological responses and might have accelerated the existing burden of mental health outcome during the COVID-19 outbreak, this burden may further increase and may stay longer depending on the time required to control the infection as well as how long will an effective vaccine really take to be available in the global market. Studies on the risk factors associated with the various mental health problems is need to be explored to manage with evidence based interventions. Some of the individual level risk factors may also get effected by the country level parameters such as countries’ policies on virus prevention at community level, healthcare infrastructure, climatic condition, concurrent burden of COVID-19 and its spreading speed, government program and policies to control the psychological responses, facilities of educational and behavioral intervention, rehabilitation centers etc. These ongoing challenges vary from one country to another country and going to be a global devastating public health crisis. Policymakers need to make effective decisions about where to focus their efforts to mitigate such burden.

### 4.2 Limitations

Around 37% of the studies were average or poor in their quality. Data were not collected based on the appropriate sampling design from a well-defined population in majority of the studies as those were collected from online survey on social media platform. Only two studies depicted the calculation of representative sample size or power assessment. Further observation of a wide heterogeneity in the reported results provides clue of lower confidence in graded evidence. Included studies are the cross-sectional studies and hence it is hard to comment on the temporal trends of mental health problems during this continuing ongoing pandemic.

## 5. Conclusion

Overall COVID-19 pandemic has been impacting on mental health of world-wide general population but frontline healthcare warriors had shown relatively having more stress, anxiety, depression and insomnia as compared to general healthcare workers and general. However, mostly these mental ailments are mild to moderate in severity. Our finding suggests that the new psychological reactions and sudden increment in burden of mental health outcomes during the COVID-19 pandemic is prompting towards the another global health emergency. Therefore a call of urgent attention and pan-region intervention are required to manage the current burden of mental health outcomes and further for future prevention.

## Data Availability

Study level data of individual studies used for meta-analysis is available with authors and will be provided on demand.

## Acknowledgements

We thank to the Department of Research, Kalinga institute of Medical Sciences, KIIT University to make available all the resources to conduct this study.

## Appendix A. Supplementary data

