## Supplementary tables for "Burden of predominant psychological reactions among the healthcare workers and general during COVID-19 pandemic phase: a systematic review and meta-analysis"

Table A1: Summary of study characteristics

| **Study** | **Country** | **Population** | **Sample**  **Size** | **Anxiety**  **Prevalence**  **(%)** | **Depression**  **Prevalence**  **(%)** | **Stress**  **Prevalence**  **(%)** | **Insomnia**  **Prevalence**  **(%)** |
| --- | --- | --- | --- | --- | --- | --- | --- |
| (Cao et al., 2020) | China | Medical Students | 7143 | 24.86 | NR | NR | NR |
| (Wang, C. et al., 2020) | China | General population | 1210 | 36.36 | 30.33 | 32.15 | NR |
| (Lai et al., 2020) | China | healthcare workers in  fever clinic and  COVID-19 wards | 1257 | 44.55 | 50.44 | 71.52 | 33.97 |
| (Wang, Y. et al., 2020) | China | General population | 600 | 6.33 | 17.17 | NR | NR |
| (Huang et al., 2020) | China | medical-staff involved  in COVID-19 | 230 | 23.04 | NR | NR | NR |
| (Qiu et al., 2020) | China,  Hong Cong,  Macau, Taiwan | General population | 52730 | NR | NR | 34.43 | NR |
| (Lu et al., 2020) | China | Medical workforce fighting  COVID-19 &  administrative staff | 2299 | 25.51^a^,  18.68^c^ | 12.1^a^,  8.17^c^ | NR | NR |
| (Nguyen et al., 2020) | Vietnam | People visiting OPD  of a hospital | 3947 | NR | 7.45 | NR | NR |
| (Zhang, W.R. et al., 2020) | China | medical as well as  non-medical health worker | 1255 | 10.45 | 10.63 | NR | 33.87 |
| (Zhang and Ma, 2020) | China | Local people | 263 | NR | NR | 7.6 | NR |
| (Ahmed et al., 2020) | China | WeChat users | 1074 | 28.96 | 37.15 | NR | NR |
| (Chen et al., 2020) | China | Paediatric medical  staff working at  high risk place | 105 | 18.1 | 29.52 | NR | NR |
| (Chew et al., 2020) | Singapore-India | health Care workers | 906 | 15.67 | 10.6 | 5.19 | NR |
| (Lei et al., 2020) | China | general population | 1593 | 8.35 | 14.69 | NR | NR |
| (Moghanibashi-Mansourieh, 2020) | Iran | general population | 10754 | 50.88 | NR | NR | NR |
| (Shacham et al., 2020) | Israel | Dentist and Dental hygienist | 338 | NR | NR | 11.54 | NR |
| (Tan et al., 2020) | China | Working Population | 673 | 6.09 | 5.94 | 3.27 | 14.56 |
| (Yuan et al., 2020) | China | General population | 939 | NR | NR | NR | 56.12 |
| (Zhu, S. et al., 2020) | China | health Care workers  frontline as well as  non-frontline | 2279 | 19.69^a^,  22^c^ | 21.25^a^,  21.69^c^ | NR | NR |
| (Yin et al., 2020) | China | Frontline healthcare workers | 371 | NR | NR | 3.78 | NR |
| (Li, J. et al., 2020) | China | general Population  (no psychiatric history)  included few COVID-cases | 5033 | 20.45 | 20.45 | NR | NR |
| (Zhou et al., 2020) | China | Adolescent | 8079 | 37.38 | 43.73 | NR | NR |
| (Liang et al., 2020) | China | young general people | 584 | NR | NR | 14.38 | NR |
| (Zhang, J. et al., 2020) | China | COVID-19 infected,  quarantined patients  and general Population | 205 | 43.88^c^, 26.31^d^,  10^e^ | 62.24^c^,  36.84^d^, 12^e^ | NR | NR |
| (Hao et al., 2020) | China | healthy subjects | 109 | 6.42 | 1.83 | 0.92 | 25.69 |
| (Jahanshahi et al., 2020) | Iran | Iranian adults | 1058 | NR | NR | 61.06 | NR |
| (Ren et al., 2020) | China | General Population | 1172 | 13.31 | 18.77 | 7.00 | 7.25 |
| (Li, X. et al., 2020) | China | frontline healthcare | 948 | NR | NR | NR | 32.81 |
| (Guo et al., 2020) | China | quarantined patients  with COVID-19 | 103 | 55.34 | 60.19 | NR | NR |
| (Gonzalez-Sanguino et al., 2020) | Spain | General Population | 3484 | 21.58 | 18.69 | 15.79 | NR |
| (Du et al., 2020) | china | frontline healthcare  workers | 310 | 9.03 | 5.48 | NR | NR |
| (Tang et al., 2020) | China | Home-quarantined  Chinese university students | 2485 | NR | 9.01 | 2.70 | NR |
| (Voitsidis et al., 2020) | Greece | elderly population | 2363 | NR | NR | NR | 37.62 |
| (Odriozola-Gonzalez et al., 2020) | Spain | students and workers  of a Spanish university | 3,707 | 35.18 | 48.10 | 40.33 | NR |
| (Abdessater et al., 2020) | France | frontline g French  urologists in training | 275 | NR | NR | 91.64 | NR |
| (Lee et al., 2020) | USA | MTurk workers | 1237 | 35.97 | 40.02 | NR | NR |
| (Liu et al., 2020) | China | frontline healthcare  workers | 512 | 12.50 | NR | NR | NR |
| (Durankus and Aksu, 2020) | Turkey | pregnant women | 260 | NR | 35.38 | NR | NR |
| (Choudhury et al., 2020) | England | frontline healthcare  workers | 106 | 60.98 | 53.85 | NR | NR |
| (Cellini et al., 2020) | Italy | General population | 1310 | 32.60 | 24.20 | 50.15 | NR |
| (Ozdin and Bayrak Ozdin, 2020) | Turkey | General Population | 343 | 45.19 | 23.62 | NR | NR |
| (Gao et al., 2020) | China | General population | 4872 | 22.60 | 48.30 | NR | NR |
| (Ozamiz-Etxebarria et al., 2020) | Spain | General population | 976 | 26.02 | 18.85 | 23.67 | NR |
| (Ni et al., 2020) | China | General population  and Health professionals | 1791 | 21.96^b^,  23.84^c^ | 19.59 | NR | NR |
| (Amerio et al., 2020) | Italy | Frontline general  practitioners | 131 | NR | 22.90 | NR | NR |
| (Zhang, C. et al., 2020) | China | frontline medical staffs | 1563 | 44.72 | 50.67 | NR | 36.08 |
| (Zhu, J. et al., 2020) | China | frontline healthcare  workers | 165 | 20.00 | 44.24 | NR | NR |
| (Mazza et al., 2020) | Italy | General population | 2766 | 18.69 | 32.75 | 27.19 | NR |
| (Consolo et al., 2020) | Italy | Dental Practitioners | 356 | 57.30 | NR | NR | NR |
| (Li, Y. et al., 2020) | China | General population | 3637 | 27.50 | 31.21 | 17.60 | 33.71 |
| (Huang and Zhao, 2020) | China | General Population and  health care workers | 7236 | 35.64^b^,  34.85^c^ | 19.82^b^,  20.21^c^ | NR | 19.82^b^,  15.76^c^ |

a- Frontline health workers; b- non-frontline health workers; c-general population; d- COVID-19 patients; e- quarantined; NR- Not Reported

Table A2: Quality assessment of included studies

| **Study** | **Sample representative** | **Sample size** | **Outcome assessment** | **Non-response rate** | **Quaity Score** |
| --- | --- | --- | --- | --- | --- |
| (Cao et al., 2020) | 1 | 1 | 2 | 1 | 5 |
| (Wang, C. et al., 2020) | 0 | 1 | 2 | 1 | 4 |
| (Lai et al., 2020) | 1 | 1 | 2 | 1 | 5 |
| (Wang, Y. et al., 2020) | 0 | 0 | 2 | 1 | 3 |
| (Huang et al., 2020) | 1 | 0 | 2 | 1 | 4 |
| (Qiu et al., 2020) | 0 | 1 | 2 | 0 | 3 |
| (Lu et al., 2020) | 1 | 1 | 2 | 1 | 5 |
| (Nguyen et al., 2020) | 1 | 1 | 2 | 1 | 5 |
| (Zhang, W.R. et al., 2020) | 0 | 1 | 2 | 0 | 3 |
| (Zhang and Ma, 2020) | 0 | 0 | 2 | 0 | 2 |
| (Ahmed et al., 2020) | 0 | 1 | 2 | 0 | 3 |
| (Chen et al., 2020) | 1 | 0 | 2 | 0 | 3 |
| (Chew et al., 2020) | 1 | 1 | 2 | 1 | 5 |
| (Lei et al., 2020) | 0 | 1 | 2 | 0 | 3 |
| (Moghanibashi-Mansourieh, 2020) | 0 | 1 | 2 | 0 | 3 |
| (Shacham et al., 2020) | 1 | 0 | 2 | 0 | 3 |
| (Tan et al., 2020) | 0 | 1 | 2 | 0 | 3 |
| (Yuan et al., 2020) | 0 | 1 | 2 | 0 | 3 |
| (Zhu, S. et al., 2020) | 0 | 1 | 2 | 0 | 3 |
| (Yin et al., 2020) | 0 | 0 | 2 | 1 | 3 |
| (Li, J. et al., 2020) | 1 | 1 | 2 | 1 | 5 |
| (Zhou et al., 2020) | 1 | 1 | 2 | 1 | 5 |
| (Liang et al., 2020) | 0 | 0 | 2 | 1 | 3 |
| (Zhang, J. et al., 2020) | 0 | 0 | 2 | 1 | 3 |
| (Hao et al., 2020) | 0 | 0 | 2 | 1 | 3 |
| (Jahanshahi et al., 2020) | 0 | 1 | 2 | 1 | 4 |
| (Ren et al., 2020) | 0 | 1 | 2 | 1 | 4 |
| (Li, X. et al., 2020) | 0 | 1 | 2 | 1 | 4 |
| (Guo et al., 2020) | 0 | 1 | 2 | 1 | 4 |
| (Gonzalez-Sanguino et al., 2020) | 1 | 1 | 2 | 1 | 5 |
| (Du et al., 2020) | 0 | 0 | 2 | 1 | 3 |
| (Tang et al., 2020) | 1 | 1 | 2 | 1 | 5 |
| (Voitsidis et al., 2020) | 0 | 1 | 2 | 1 | 4 |
| (Odriozola-Gonzalez et al., 2020) | 0 | 1 | 2 | 1 | 4 |
| (Abdessater et al., 2020) | 1 | 1 | 2 | 1 | 5 |
| (Lee et al., 2020) | 0 | 1 | 2 | 1 | 4 |
| (Liu et al., 2020) | 1 | 1 | 2 | 1 | 5 |
| (Durankus and Aksu, 2020) | 0 | 0 | 2 | 1 | 3 |
| (Choudhury et al., 2020) | 0 | 0 | 2 | 1 | 3 |
| (Cellini et al., 2020) | 0 | 1 | 2 | 1 | 4 |
| (Ozdin and Bayrak Ozdin, 2020) | 0 | 1 | 2 | 1 | 4 |
| (Gao et al., 2020) | 0 | 1 | 2 | 1 | 4 |
| (Ozamiz-Etxebarria et al., 2020) | 0 | 1 | 2 | 1 | 4 |
| (Ni et al., 2020) | 0 | 1 | 2 | 1 | 4 |
| (Amerio et al., 2020) | 0 | 1 | 2 | 1 | 4 |
| (Zhang, C. et al., 2020) | 0 | 1 | 2 | 1 | 4 |
| (Zhu, J. et al., 2020) | 1 | 1 | 2 | 1 | 5 |
| (Mazza et al., 2020) | 0 | 1 | 2 | 1 | 4 |
| (Consolo et al., 2020) | 0 | 1 | 2 | 1 | 4 |
| (Li, Y. et al., 2020) | 0 | 1 | 2 | 1 | 4 |
| (Huang and Zhao, 2020) | 0 | 1 | 2 | 1 | 4 |

Table A3: Assessment of Confidence in our reported finding by GRADE approach

| **№ of studies** | **Certainty assessment** | | | | | | | | | | | | | | | **Effect** | | | | | | | **Certainty** | **Importance** |
| --- | --- | --- | --- | --- | --- | --- | --- | --- | --- | --- | --- | --- | --- | --- | --- | --- | --- | --- | --- | --- | --- | --- | --- | --- |
|  | **Study design** | | | **Risk of bias** | | **Inconsistency** | | | | | **Indirectness** | | | **Imprecision** | **Other considerations** | **№ of events** | | | **№ of individuals** | **Rate (95% CI)** | | |  |  |
| Anxiety | | | | | | | | | | | | | | | | | | | | | | | | |
| 43 | observational studies | | | serious ^a^ | | very serious ^b^ | | | | | not serious | | | not serious | publication bias strongly suspected strong association all plausible residual confounding would reduce the demonstrated effect ^c^ | 27525 | | | 90080 | event rate 26.59 per 100 (22.81 to 30.36) | | ⨁⨁◯◯ LOW | | IMPORTANT |
| Anxiety among Frontline health care Workers | | | | | | | | | | | | | | | | | | | | | | | | |
| 10 | observational studies | | | serious ^a^ | | very serious ^b^ | | | | | not serious | | | not serious | strong association all plausible residual confounding would reduce the demonstrated effect | 2065 | | | 6545 | event rate 27.2 per 100 (18.1 to 36.31) | | ⨁⨁⨁◯ MODERATE | | IMPORTANT |
| Anxiety among Second line health care worker | | | | | | | | | | | | | | | | | | | | | | | | |
| 7 | observational studies | | | serious ^a^ | | very serious ^b^ | | | | | not serious | | | not serious | strong association all plausible residual confounding would reduce the demonstrated effect | 3199 | | | 12124 | event rate 26.89 per 100 (20.35 to 33.43) | | ⨁⨁⨁◯ MODERATE | | IMPORTANT |
| Anxiety among general population | | | | | | | | | | | | | | | | | | | | | | | | |
| 24 | observational studies | | | serious ^a^ | | very serious ^b^ | | | | | not serious | | | not serious | publication bias strongly suspected strong association all plausible residual confounding would reduce the demonstrated effect ^c^ | 21778 | | | 68742 | event rate 25.87 per 100 (20.51 to 31.24) | | ⨁⨁◯◯ LOW | | IMPORTANT |
| Mild Anxiety | | | | | | | | | | | | | | | | | | | | | | | | |
| 18 | observational studies | | | serious ^a^ | | very serious ^b^ | | | | | not serious | | | not serious | publication bias strongly suspected strong association all plausible residual confounding would reduce the demonstrated effect ^c^ |  | | |  | event rate 16.69 per 100 (12.27 to 21.11) | | ⨁⨁◯◯ LOW | | NOT IMPORTANT |
| Moderate Anxiety | | | | | | | | | | | | | | | | | | | | | | | | |
| 18 | observational studies | | | serious ^a^ | | very serious ^b^ | | | | | not serious | | | not serious | publication bias strongly suspected all plausible residual confounding would reduce the demonstrated effect ^c^ |  | | |  | event rate 7.34 per 100 (4.4 to 10.27) | | ⨁◯◯◯ VERY LOW | | IMPORTANT |
| Severe Anxiety | | | | | | | | | | | | | | | | | | | | | | | | |
| 17 | observational studies | | | serious ^a^ | | very serious ^b^ | | | | | not serious | | | not serious | publication bias strongly suspected all plausible residual confounding would reduce the demonstrated effect ^c^ |  | | |  | event rate 5.39 per 100 (3.19 to 7.59) | | ⨁◯◯◯ VERY LOW | | CRITICAL |
| Depression | | | | | | | | | | | | | | | | | | | | | | | | |
| 43 | observational studies | | | serious ^d^ | | very serious ^b^ | | | not serious | | | | | not serious | publication bias strongly suspected strong association all plausible residual confounding would reduce the demonstrated effect ^c^ | | | 21019 | 77932 | event rate 26.17 per 100 (21.84 to 30.5) | | ⨁⨁◯◯ LOW | | CRITICAL |
| Depression among Frontline health care worker | | | | | | | | | | | | | | | | | | | | | | | | |
| 9 | observational studies | | | serious ^a^ | | very serious ^b^ | | | not serious | | | | | not serious | strong association all plausible residual confounding would reduce the demonstrated effect | | 1927 | | 5958 | event rate 32.1 per 100 (17.97 to 46.22) | | ⨁⨁⨁◯ MODERATE | | CRITICAL |
| Depression among Second line health Care Workers | | | | | | | | | | | | | | | | | | | | | | | | |
| 5 | observational studies | | | serious ^a^ | | very serious ^b^ | | | not serious | | | | | not serious | strong association all plausible residual confounding would reduce the demonstrated effect | | 815 | | 4625 | event rate 15.72 per 100 (11.28 to 20.15) | | ⨁⨁⨁◯ MODERATE | | CRITICAL |
| Depression among general Population | | | | | | | | | | | | | | | | | | | | | | | | |
| 25 | observational studies | | | serious ^a^ | | very serious ^b^ | | | not serious | | | | | not serious | publication bias strongly suspected strong association all plausible residual confounding would reduce the demonstrated effect ^c^ | | 17496 | | 62195 | event rate 25.87 per 100 (20.22 to 31.53) | | ⨁⨁◯◯ LOW | | CRITICAL |
| Mild Depression | | | | | | | | | | | | | | | | | | | | | | | | |
| 12 | observational studies | | | serious ^a^ | | very serious ^b^ | | | not serious | | | | | not serious | publication bias strongly suspected strong association all plausible residual confounding would reduce the demonstrated effect ^c^ | |  | |  | event rate 20.81 per 100 (13.94 to 27.69) | | ⨁⨁◯◯ LOW | | NOT IMPORTANT |
| Moderate Depression | | | | | | | | | | | | | | | | | | | | | | | | |
| 13 | observational studies | | | serious ^a^ | | very serious ^b^ | | | not serious | | | | | not serious | publication bias strongly suspected all plausible residual confounding would reduce the demonstrated effect ^c^ | |  | |  | event rate 7.41 per 100 (4.93 to 9.9) | | ⨁◯◯◯ VERY LOW | | IMPORTANT |
| Severe depression | | | | | | | | | | | | | | | | | | | | | | | | |
| 12 | observational studies | | | serious ^a^ | | very serious ^b^ | | | not serious | | | | | not serious | publication bias strongly suspected all plausible residual confounding would reduce the demonstrated effect ^c^ | |  | |  | event rate 4.61 per 100 (2.65 to 6.58) | | ⨁◯◯◯ VERY LOW | | CRITICAL |
| Stress | | | | | | | | | | | | | | | | | | | | | | | | |
| 20 | observational studies | | | serious ^a^ | | | | very serious ^b^ | | serious ^d^ | | | | not serious | strong association all plausible residual confounding would reduce the demonstrated effect | | 25210 | | 79138 | | event rate 26.16 per 100 (17.73 to 34.59) | ⨁⨁◯◯ LOW | | IMPORTANT |
| Stress among frontline health care worker | | | | | | | | | | | | | | | | | | | | | | | | |
| 3 | observational studies | | | serious ^e^ | | | | very serious ^b^ | | | | serious ^d^ | | serious ^c^ | strong association all plausible residual confounding would reduce the demonstrated effect | | 1165 | | 1903 | | event rate 55.63 0.36 110.9 per 100 (0.36 to 100) | ⨁◯◯◯ VERY LOW | | IMPORTANT |
| Stress among Second line health care worker | | | | | | | | | | | | | | | | | | | | | | | | |
| 3 | observational studies | | | serious ^a^ | | | | very serious ^b^ | | | | serious ^d^ | | serious ^c^ | all plausible residual confounding would reduce the demonstrated effect | | 86 | | 1244 | | event rate 7.01 per 100 (3.11 to 10.92) | ⨁◯◯◯ VERY LOW | | IMPORTANT |
| Stress among general population | | | | | | | | | | | | | | | | | | | | | | | | |
| 13 | observational studies | | | serious ^a^ | | | | very serious ^b^ | | | | serious ^d^ | | not serious | strong association all plausible residual confounding would reduce the demonstrated effect | | 23892 | | 73506 | | event rate 25.54 per 100 (17.8 to 33.29) | ⨁⨁◯◯ LOW | | IMPORTANT |
| Insomnia | | | | | | | | | | | | | | | | | | | | | | | | |
| 11 | observational studies | | | | serious ^a^ | | very serious ^b^ | | | | | | not serious | not serious | strong association all plausible residual confounding would reduce the demonstrated effect | | 7490 | | 27775 | event rate 32.87 per 100 (23.94 to 41.8) | | ⨁⨁⨁◯ MODERATE | | IMPORTANT |
| Insomnia among frontline health care worker | | | | | | | | | | | | | | | | | | | | | | | | |
| 3 | | observational studies | | | serious ^a^ | | serious ^b^ | | | | | | not serious | serious ^d^ | publication bias strongly suspected strong association ^c^ | | 1362 | | 3828 | event rate 34.44 per 100 (32.54 to 36.34) | | ⨁◯◯◯ VERY LOW | | IMPORTANT |
| Insomnia among second line health care worker | | | | | | | | | | | | | | | | | | | | | | | | |
| 2 | | | observational studies | | very serious ^b,d^ | | serious ^b^ | | | | | | not serious | serious ^d^ | strong association all plausible residual confounding would reduce the demonstrated effect | | 1270 | | 3505 | event rate 33.96 per 100 (32.48 to 35.43) | | ⨁⨁◯◯ LOW | | IMPORTANT |
| Insomnia among general population | | | | | | | | | | | | | | | | | | | | | | | | |
| 6 | | | observational studies | | serious ^b^ | | serious ^b^ | | | | | | not serious | not serious | strong association all plausible residual confounding would reduce the demonstrated effect dose response gradient | | 4858 | | 20442 | event rate 29.36 per 100 (17.02 to 41.69) | | ⨁⨁⨁⨁ HIGH | | IMPORTANT |

a. Non-random sampling was used for most of the studies; b. Heterogeneity index measured using I2 statistic is very high; c. Few Number of studies; d. There was a wide variety in the scales and definition of stress among studies; e. The quality assessed by New Castel Ottawa scale is poor
